# Automatic Detection of Cardiac Conditions from Photos of Electrocardiogram (ECG) Captured by Smartphones

**DOI:** 10.1101/2023.11.01.23297947

**Authors:** Chun-Ka Wong, Yuk-Ming Lau, Hin-Wai Lui, Wai-Fung Chan, Wing-Chun San, Mi Zhou, Yangyang Cheng, Duo Huang, Wing-Hon Lai, Yee-Man Lau, Chung-Wah Siu

**Author notes:** These authors are co-first authors (Chun-Ka Wong, Yuk-Ming Lau, Hin-Wai Lui). These authors are co-corresponding authors (Chun-Ka Wong, Chung-Wah Siu). **Correspondence Address: Chun-Ka WONG, MBBS**, Clinical Assistant Professor, Department of Medicine, The University of Hong Kong, Address: Rm 1919, Block K, Queen Mary Hospital, Hong Kong SAR, **Chung-Wah SIU, MD**, Honorary Clinical Professor, Department of Medicine, The University of Hong Kong, Address: Rm 1919, Block K, Queen Mary Hospital, Hong Kong SAR.

## Abstract

**Background:** Artificial intelligent electrocardiogram (ECG) diagnostic algorithms can achieve cardiologist-level accuracy, but their clinical use is limited as they cannot be installed in older ECG machines.

**Objective:** To develop a smartphone application that extracts and analyzes ECG waveforms from photos using machine learning techniques.

**Methods:** A smartphone application was developed to automatically extract ECG waveforms from photos taken by clinicians using computer vision and machine learning. Custom designed machine learning models were developed to perform waveform identification, gridline removal, and scale calibration. The extracted voltage-time series waveforms were analyzed using a pre-trained machine learning-based diagnostic algorithms, and the accuracy of the proof-of-concept application was assessed.

**Results:** Waveforms from 40,516 scanned and 444 photographed ECGs were automatically extracted. 12,828 of 13,258 (96.8%) scanned and 5,399 of 5,743 (94.0%) photographed waveforms were correctly cropped and labelled. 11,604 of 12,735 (91.1%) scanned and 5,062 of 5,752 (88.0%) photographed waveforms achieved successful voltage-time signal extraction after automatic gridline and background noise removal. The AF diagnostic algorithm achieved 91.3% sensitivity, 94.2% specificity, 95.6% positive predictive value, 88.6% negative predictive value and 93.4% F1 score.

**Conclusion:** Using computer vision and machine learning techniques to detect cardiac conditions from photos of ECGs taken with smartphones is feasible. This platform can enable widespread deployment of the latest machine learning-based ECG diagnostic algorithms.

## INTRODUCTION

In recent years, rapid advances in artificial neural network technologies have revolutionized all industries including the cardiology field. ^1–3^ Increasingly accurate electrocardiogram (ECG) diagnostic algorithms for detecting conduction system disorders, ^4–6^ myocardial infarction, ^7,8^ valvular disease, ^9–11^ cardiomyopathies, ^12,13^ heart failure with reduced ejection fraction, ^14^ electrolyte disturbance, ^15^ and accessory pathways, ^16^ and for predicting mortality risk ^17^ have been developed by researchers using machine learning approaches. However, the utilization of these diagnostic algorithms in real-world clinical practice is hindered because most existing ECG machines cannot install and apply these latest algorithms.

A cost-effective way to enable global adoption of these latest ECG diagnostic algorithms is to utilize smartphones as ECG interpreters. ECG images, either scanned or photographed, cannot be directly analyzed by ECG diagnostic algorithms due to file format mismatch. Instead of an image, an ECG has to be converted to waveform data in voltage-time series format before being analyzed. ^4–16^ A system that can automatically extract ECG waveforms from photos and convert them to voltage-time series format is needed to apply ECG diagnostic algorithms to ECG images. Such system can enable automatic detection of cardiac conditions from photographed ECGs captured by smartphones.

Previous attempts to extract waveforms from scanned ECG by computer vision-based software often failed to achieve fully-automatic waveform extraction and required extensive manual intervention.^18–22^ Recent advances in artificial intelligent (AI) object detection and image segmentation techniques have made it possible to achieve fully automatic waveform extraction from photos of ECGs taken using a smartphone.

The objective of this study was to develop and validate DigitHeart, a smartphone application that employs computer vision and machine learning techniques to automatically extract ECG waveforms from ECG images. The extracted waveforms in voltage-time series format were further analyzed by machine learning-based diagnostic algorithms. The accuracies of waveform extraction and diagnostic algorithm were evaluated.

## METHODS

The study protocol was approved by the Institutional Review Board of the University of Hong Kong/Hospital Authority Hong Kong West Cluster (UW 12-177) and complied with the Declaration of Helsinki.

### Raw dataset

Patients with atrial fibrillation (AF) were drawn from the AF registry of Queen Mary Hospital, University of Hong Kong and included in this study. Since this study was based on anonymized data from a hospital AF registry, need for patient consent was waived. Scanned grayscale ECGs and photographed color ECGs were acquired. All ECGs were anonymized by masking the patient’s identity before use. Clinical data of patients were acquired using the Clinical Data Analysis and Reporting System (CDARS) of the Hospital Authority, Hong Kong.

### Training dataset

Machine learning models were trained using a paired training dataset with raw and annotated images. ECG waveforms were extracted from the raw images using computer vision techniques, which will be described in detail in the following sections. A web-based graphic user interface was used to manually review and edit the extracted waveforms to enhance accuracy. Django 2·2·1 was used as server to store all raw and annotated datasets.

### Smartphone application and deep learning models for ECG waveform extraction

The DigitHeart system is a cloud-based smartphone application for automatically extracting waveforms from images of ECGs and analyzing them using machine learning-based ECG diagnostic algorithms. To use the DigitHeart system, clinicians need to photograph ECGs using the smartphone application which will then upload them to the server for further processing. The backend server processes the images in three steps. (Figure 1, Supplementary Table S1) Firstly, an object detection model was used to locate, and label waveforms on ECGs. Secondly, an image segmentation model removed gridlines and other background noise. Finally, a novel scale calibration technique was applied to the voltage marker to derive the voltage-time series data. Then the ECG waveforms can be extracted for further analysis.

**Figure 1.**
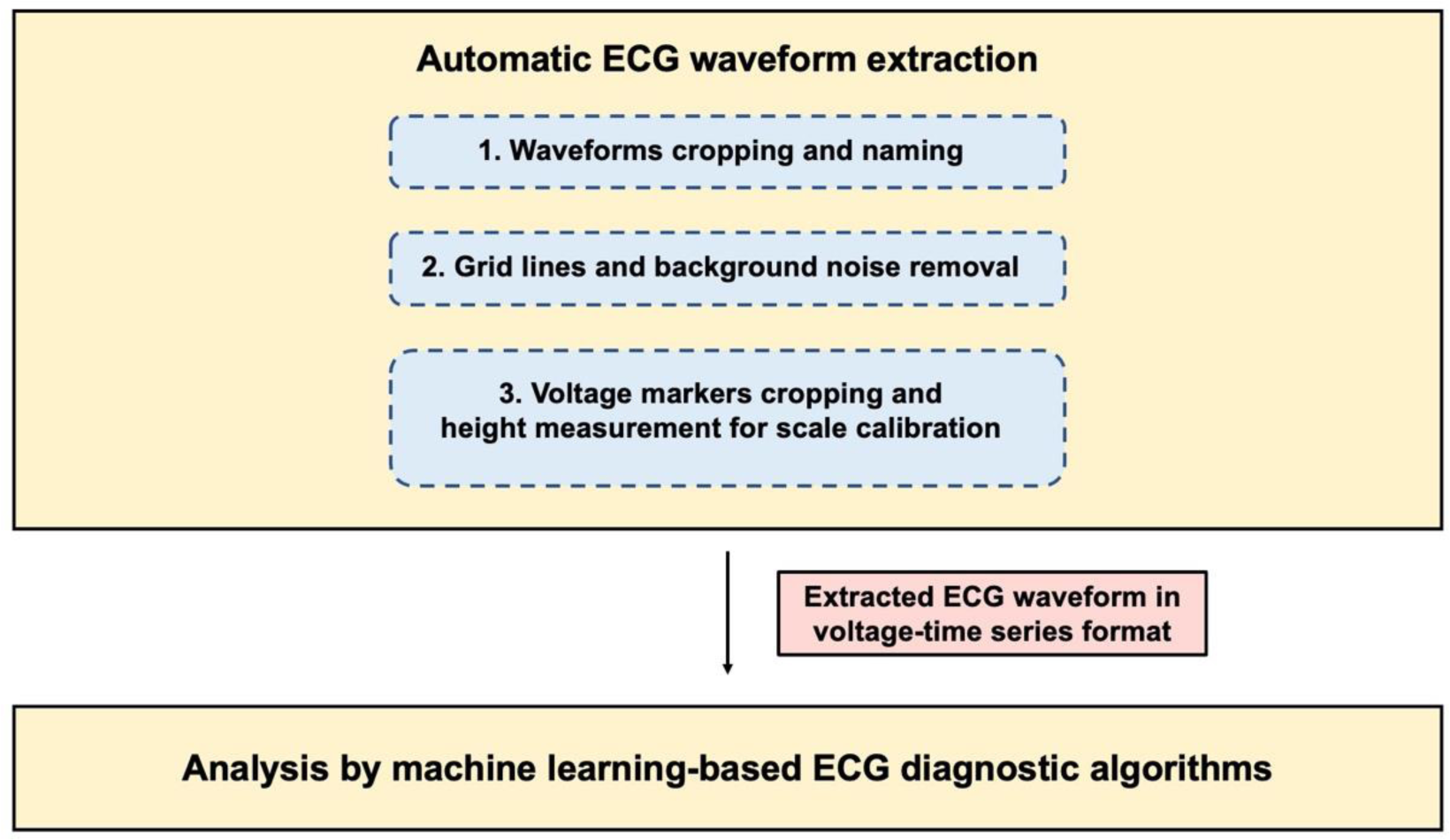
DigitHeart is an artificial intelligent powered system trained for fully automatic electrocardiogram (ECG) waveform extraction from scanned and photographed ECGs. It consists of three steps. Firstly, an object detection model was used to locate, and label waveforms on ECGs. Secondly, an image segmentation model removed gridlines and other background noise. Finally, a novel scale calibration technique was applied to the voltage marker to derive the voltage-time series data. Then the ECG waveforms can be automatically extracted for further analysis.

### Direct diagnosis derivation from photos of ECGs

The digitalized ECG waveforms are versatile and can fit into most existing diagnostic algorithms. A proof-of-concept application to detect AF in ECG was performed in this study by using a previously trained AF classifier from our group.^7^ The accuracy of the diagnostic algorithm to diagnose AF was evaluated using a dataset with cardiac rhythm labelled by cardiologists as the gold standard.

### Statistical analysis

Normal and discrete variables are presented as mean ± standard deviation and percentages, respectively. Sensitivity, specificity, positive predictive value, negative predictive value, and F1 score, were calculated for the AF classifier to assess their performance.

## RESULTS

### Data characteristics

10,945 AF patients from Queen Mary Hospital, Hong Kong, of whom 5,469 (50·0%) and 5,476 (50·0%) being male and female respectively, were included in this study. At the time of data acquisition, their mean age was 80·4 ± 11·1 years. 40,516 grayscale scanned ECGs and 444 color photographed ECGs were included. (Figure 2)

**Figure 2.**
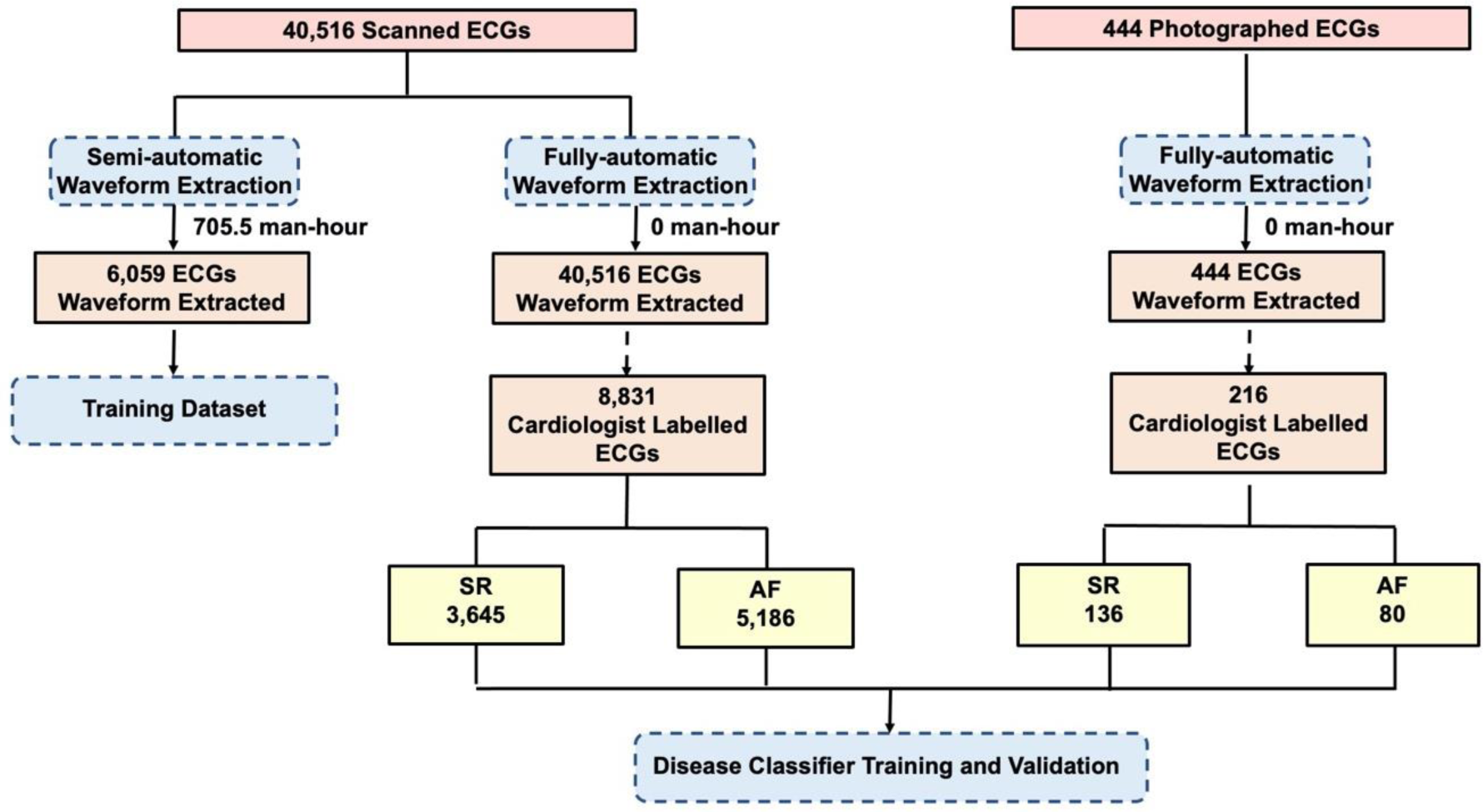
To generate a training dataset, waveforms from 6,059 scanned electrocardiograms (ECGs) were extracted using computer vision techniques. A web-based graphic user interface was used to manually review and edit the extracted waveforms to enhance accuracy. 8,831 randomly selected scanned ECGs and 216 color photographed ECGs were reviewed by cardiologists who assigned one of two possible labels to the observed cardiac rhythm: sinus rhythm (SR), and atrial fibrillation (AF).

### Waveform localization, and labelling

To generate a training dataset, technicians manually labelled all waveforms on 6,059 scanned ECGs using our web-based data annotation platform. (Figure 3A) To compensate for limited sample size, the manually generated dataset was augmented by random rotations, brightness and contrast adjustment, and noise addition. Hence each image had several augmented versions. These increased the effective training samples and helped to reduce overfitting during neural network training. (Figure 3B)

**Figure 3.**
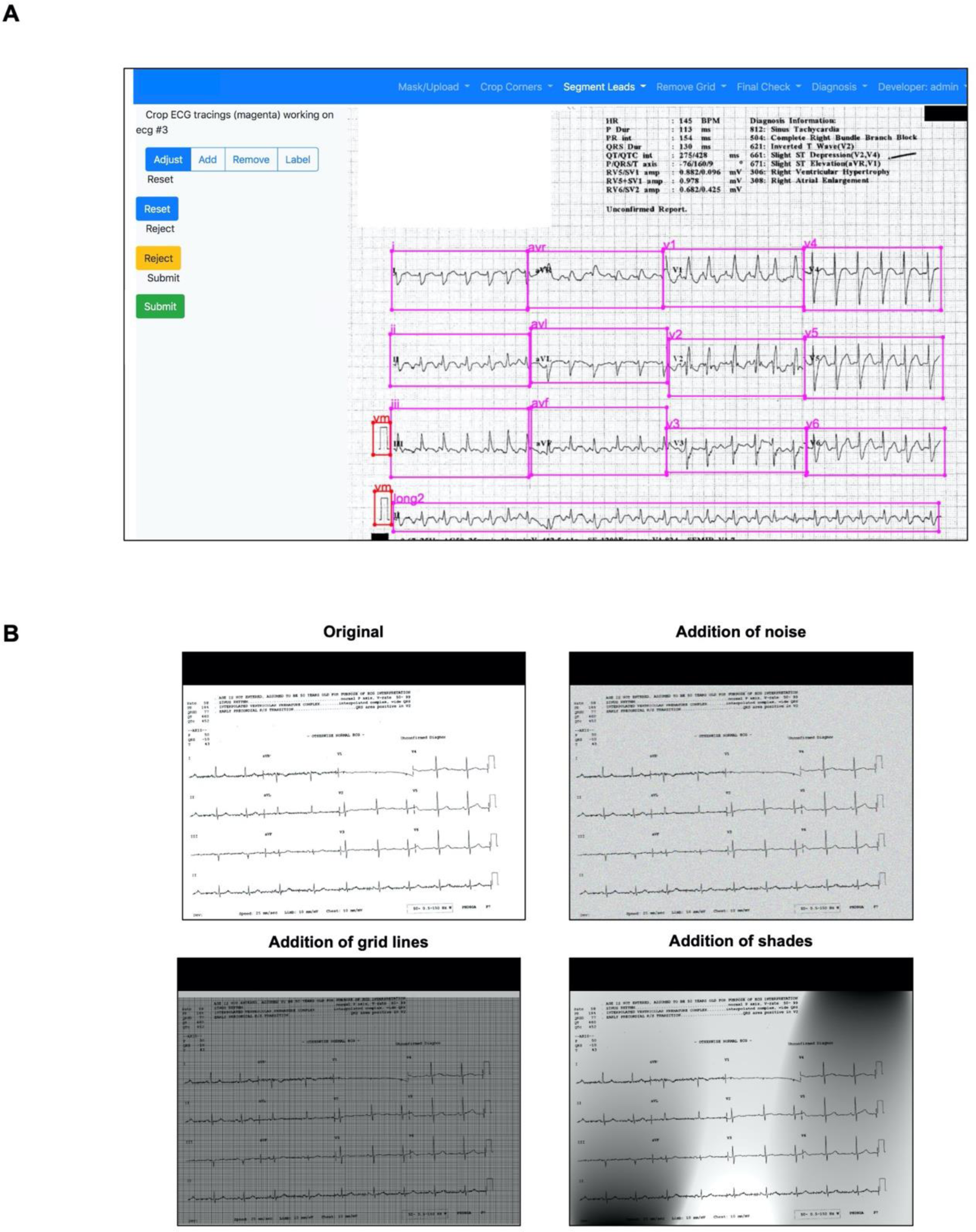
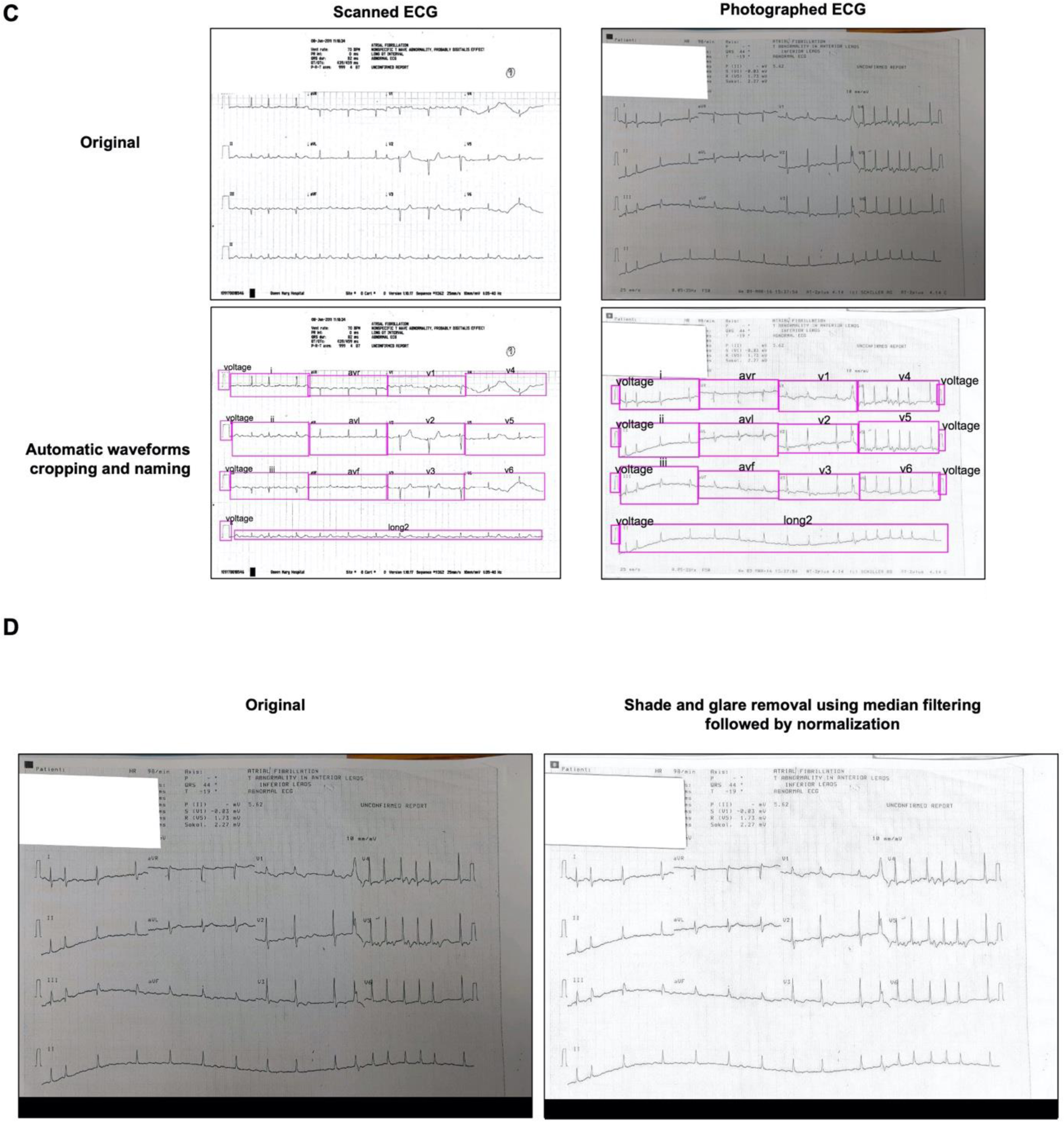
Waveforms localization, cropping and naming. (A) To generate a training dataset, technicians manually labelled all waveforms on 6,059 scanned electrocardiograms (ECGs) using our web-based data annotation platform. (B) The manually generated dataset was augmented by random rotations, brightness and contrast adjustment, and noise addition to increase the effective training samples to enhance neural network training and reduce overfitting. (C) Using the augmented dataset, an object detection neural network was trained to automatically locate and name ECG lead segments using Tensorflow GPU version 1·15 (Alphabet Inc, California, USA) and Tensorflow Object Detection API (Alphabet Inc, California, USA). The model used Faster R-CNN with Inception V2 as the backbone. (D) Photographed electrocardiograms (ECGs) had sub-optimal lighting compared with scanned ECGs due to shading and glare that affected waveform extraction. Local background subtraction with median filtering using a large-sized filter was applied followed by normalization using OpenCV 4·1·0·25 to remove shading and glare.

An object detection neural network was trained using the augmented dataset to locate and label ECG lead segments. Tensorflow GPU version 1·15 (Alphabet Inc, California, USA) and Tensorflow Object Detection API (Alphabet Inc, California, USA) were used for this purpose. The model used Faster R-CNN with Inception V2 as the backbone. It was trained with a momentum optimizer and a progressive declining learning rate from 2×10^−4^ to 1×10^−7^. Randomly selected 943 scanned and all 444 photographed ECGs were used to assess performance of automatic waveform localization, and labelling. (Figure 3C) In order to minimize the effect of suboptimal lighting due to shading and glare, photographed ECGs underwent local background subtraction with median filtering using a large-sized filter followed by normalization using OpenCV 4·1·0·25 before processed by the objection detection model. (Figure 3D). As a standard 12 leads ECG contains multiple leads, a total of 13,258 scanned leads and 5,743 photographed leads were evaluated. Among the selected scanned and photographed ECGs, 12,828 of 13,258 (96·8%) and 5,399 of 5,743 (94·0%) waveforms, respectively were correctly localized, and labelled. (Figure 4)

**Figure 4.**
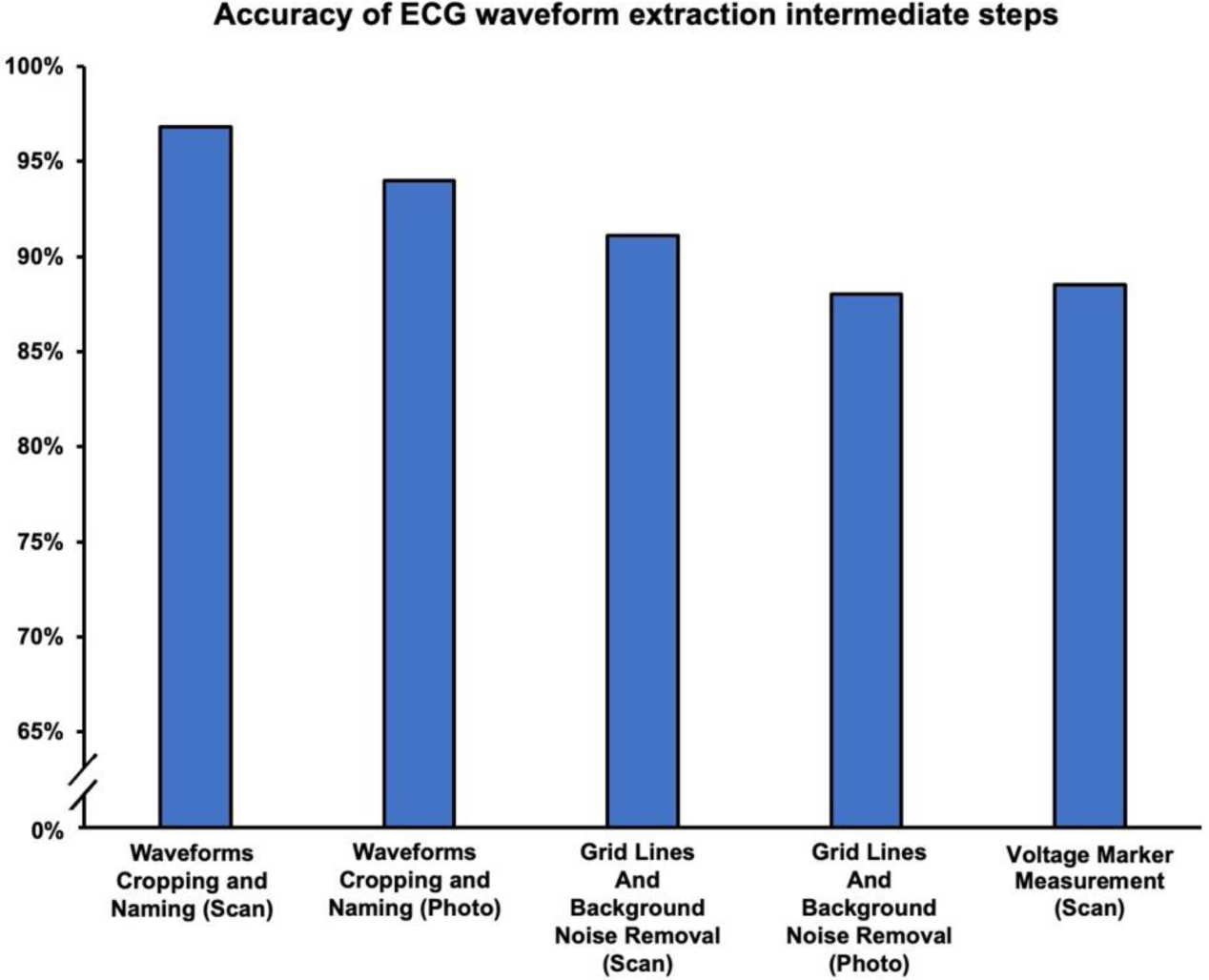
943 randomly selected scanned electrocardiograms (ECGs) and all 444 photographed ECGs were selected to assess the performance of fully automatic waveforms extraction. Among the selected scanned ECGs and photographed ECGs, 12,828 of 13,258 (96·8%) and 5,399 of 5,743 (94·0%) waveforms were correctly locate and labelled, 11,604 of 12,735 (91·1%) and 5,062 of 5,752 (88·0%) waveforms achieved successful grid and background noise removal, and 801 of 905 (88·5%) scanned ECGs had voltage marker height correctly measured.

### Gridlines and background noise removal

Gridlines and background noise had to be removed thoroughly before extracting ECG waveform signals. To generate training dataset for waveform extraction, a computer vision-based system was created using OpenCV 4·1·0·25 to remove gridlines. Since ECG tracings were darker than the background gridlines, binary thresholding was used to filter out the gridlines. However, the scanned ECGs were acquired from multiple hospitals in Hong Kong with consequent huge variation in scanning quality, brightness, and contrast. It was not possible to use the same threshold value to remove gridlines from all ECGs. To address this, multiple threshold values were applied to each ECG. The technicians manually evaluated different threshold values of the ECG images and selected the version with the least remaining gridlines and most preserved ECG signals. (Figure 5A)

**Figure 5.**
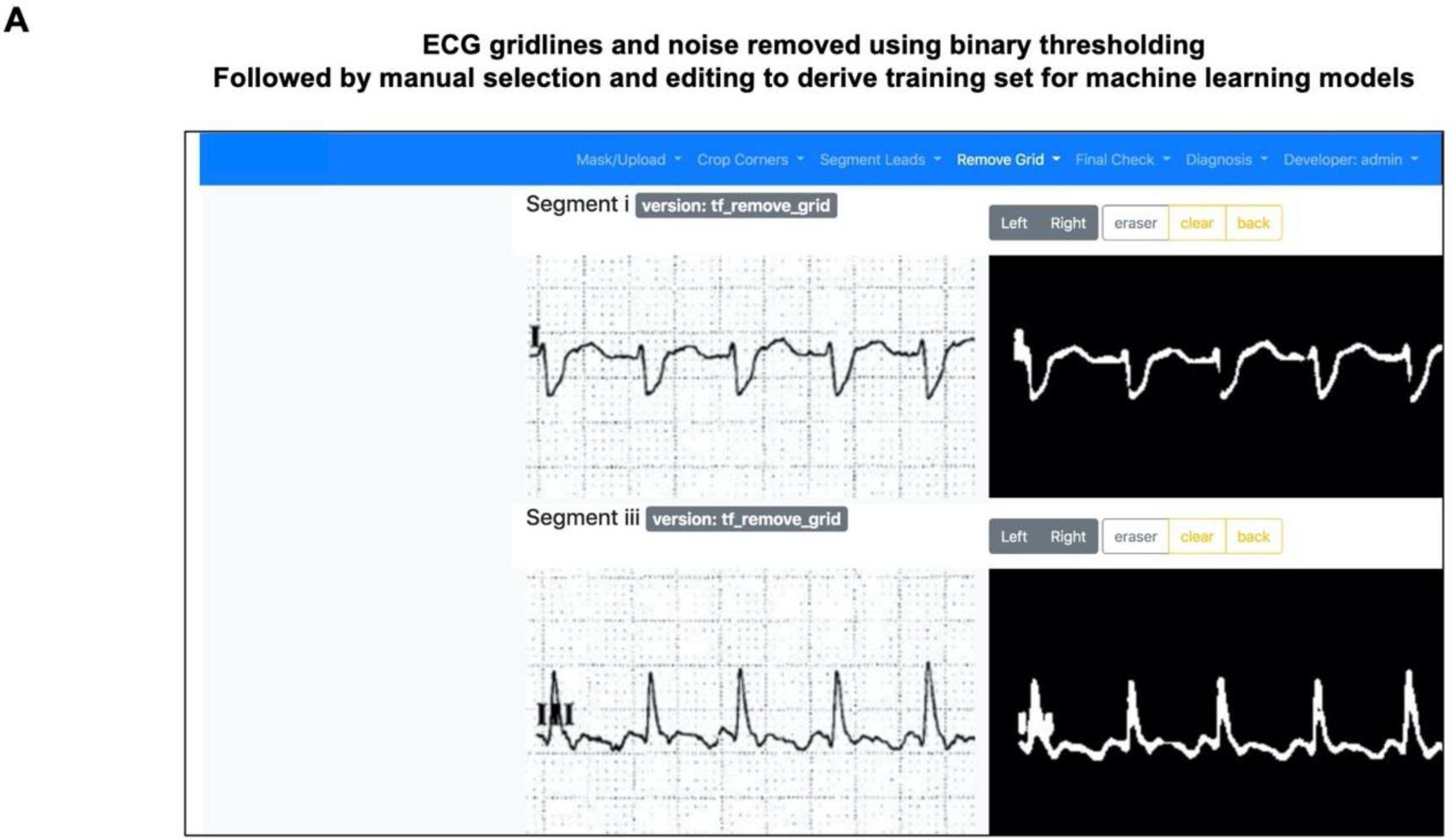
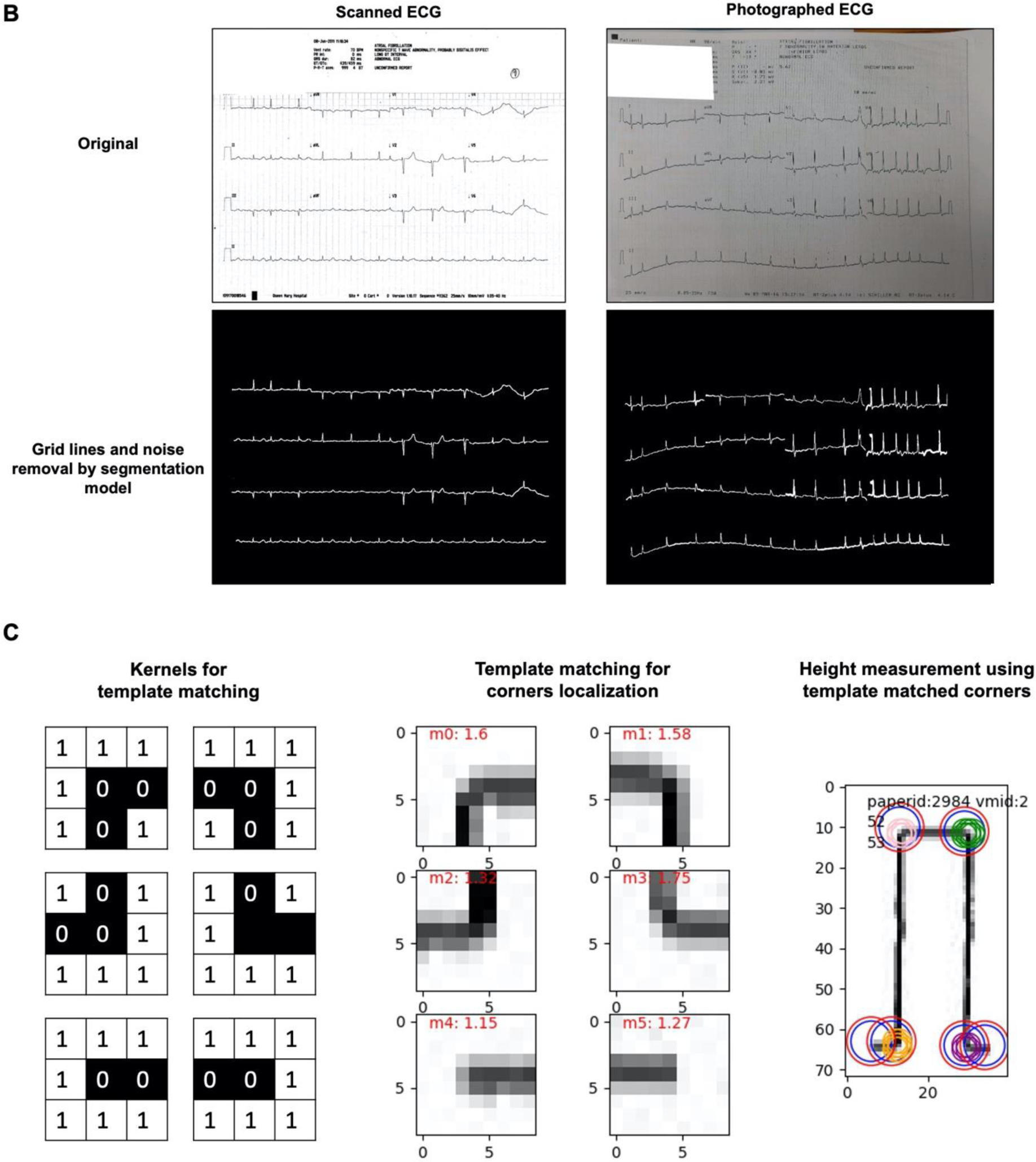
Gridlines and background noise removal (A) Since ECG tracings were darker in color than the background gridlines, binary thresholding using OpenCV 4.1.0.25 was used to filter out the gridlines. Multiple threshold values were applied to each ECG. Technicians manually evaluated different threshold values of the ECG images and selected the version with the least remaining gridlines and most preserved ECG signals. (B) An image segmentation model with a UNet architecture and Resnet101 as backbone was trained using the dataset prepared to automatically remove gridlines and background noise. (C) Voltage markers for scale calibration. The height of voltage marker was measured using template matching with OpenCV 4·1·0·25. Briefly, six kernels with size 3 x 3 pixels were used to represent six features of the voltage marker, including the four corners and the two blind-ended sides. Each kernel was convolved over the entire image. The location with the minimal sum of absolute differences was defined as the most likely location of the six representative features. The height of voltage markers in pixel-units was measured accordingly. Voltage markers had a fixed height of 10mm on standard ECGs. By measuring the height and width of the voltage markers in the images, it was possible to convert pixel-units to voltage-units and time-unit using conversion ratios of 10mm/mV, and 25mm/s.

A training set was created by pairing the original ECG images with the ‘cleaned’ version created using the aforementioned technique. Data augmentation, such as random rotations, brightness and contrast adjustment, and noise addition, were implemented to increase the effective training size. An UNet architecture image segmentation model with Resnet101 as feature extractor was used for training. ^23^ Adam optimizer with an initial learning rate of 1×10^−5^ was used. Among 943 scanned and 444 photographed ECGs reviewed, successful gridlines and background noise removal were successful in 11,604 of 12,735 (91·1%) and 5,062 of 5,752 (88·0%) lead segments respectively. (Figures 4 and 5B)

### Voltage markers for scale calibration

In the previous steps, ECG waveforms were extracted in pixel-units. These signals had to be converted to voltage-units and time-units before they could be analyzed by ECG diagnostic algorithms. A novel approach was developed to achieve scale calibration using voltage markers printed on ECGs. Voltage markers had a fixed height of 10mm on standard ECGs. By measuring the height of the voltage markers in the images, it was possible to convert pixel-units to voltage-units and time-unit using conversion ratios of 10mm/mV, and 25mm/s.

To create a training set for the object detection model to locate the voltage markers, technicians manually labelled the markers on the ECGs via the web application. (Figure 3A) The same waveform localization, and labelling model was trained additionally using these data to localize the voltage markers. (Figure 3C) After obtaining the voltage markers, their height was automatically measured using a template matching technique in OpenCV 4·1·0·25. Briefly, six kernels with size 3 x 3 pixels were used to represent six features of the voltage marker, including the four corners and the two blind-ended sides. Each kernel was convolved over the entire image. The location with the minimal sum of absolute differences was defined as the most likely location of the six representative features. The height of the voltage markers in pixel-units was measured accordingly. (Figure 5C) The height of each voltage marker was 10mm on a printed ECG by default. By measuring the height and width of the voltage markers in the images, it was possible to convert pixel-units to voltage-units and time-unit using conversion ratios of 10mm/mV, and 25mm/s. Using this combined approach, 801 of 905 (88·5%) scanned ECGs had voltage marker heights correctly measured. (Figure 4)

### Human resources utilization

In the initial phase during which ECG waveforms were manually extracted by technicians aided by computer vision techniques, 6,059 scanned ECGs were processed over 705·5 man-hours (8·59 ECGs per man-hour). The fully autonomous DigitHeart system improved the efficiency of ECG digitization. Waveforms from 40,516 scanned and 444 photographed ECGs were extracted automatically, which saved about 4,768 man-hours. (Figure 2)

### Direct diagnosis derivation from photos of ECGs

A proof-of-concept smartphone application was developed to demonstrate that the ECG waveforms extracted using DigitHeart could be utilized to directly derive a clinical diagnosis. (Supplementary Figure S1 and Video 1) As a photographed ECG has distorted geometry and is not perfectly rectangular, geometry transformation was performed using Canny edge detector, SmartCropper v1·2·5, and Pillow 8·2·0, to convert the photos to a rectangular shape with aspect ratio of A4 paper (210:297) before further processing. (Supplementary Figure S1B) Information related to patient’s identity was masked before the image was sent to server for analysis. (Supplementary Figures S1C, S1D, and S1E)

8,831 randomly selected scanned ECGs and 216 color photographed ECGs were reviewed by cardiologists who assigned one of two possible labels to the observed cardiac rhythm: sinus rhythm, and AF. Diagnosis of AF was established if there were no discernible repeating P waves or irregular RR intervals. 5,186 (58·7%), and 3,645 (41·3%) scanned ECGs were labelled as AF, and sinus rhythm respectively. 80 (37%), and 136 (63%) photographed ECGs were labelled as AF, and sinus rhythm respectively. (Supplementary Table 1)

An AF classifier, which combined both convolutional and recurrent neural networks based on a previous method from our group, was used to analyze the selected ECGs. The AF classifier was able to correctly classify 4,807 and 3,561 ECG as AF, and sinus rhythm respectively. It achieved 91·3% sensitivity, 94·2% specificity, 95·6% positive predictive value, 88·6% negative predictive value and 93·4% F1 score. (Supplementary Table 2)

## DISCUSSION

With the rapid advancement in artificial intelligence, increasingly accurate diagnostic algorithms for 12-lead ECGs have been developed by researchers. These algorithms can identify a wide range of cardiac conditions and predict clinical outcomes. ^4,5,7–10,12–15^ However, it is challenging to deploy these systems on a global scale because most ECG machines currently in use do not enable installation of new diagnostic algorithms. Replacing all ECG machines with newer generation machines with machine learning-based diagnostic algorithms is also impractical from a resource perspective. A cost-effective solution to enable wide adoption of these newer generation ECG diagnostic algorithms can be achieved using smartphones as ECG interpreters. A system which enables automatic extraction of ECG waveforms from photos, conversion to voltage-time format, and analysis using machine learning-based diagnostic algorithms, is required. The present study demonstrated feasibility of automatically extracting ECG waveforms from photos using custom designed machine learning models to perform waveform identification, gridline removal, and scale calibration, and achieving high accuracy.

In the past, specialized software was developed using traditional computer vision techniques to extract ECG waveforms from scanned ECGs. Methods such as Otsu’s algorithm and other thresholding methods were able to achieve grid and background noise removal with modest efficacy ^18–22^. Nonetheless these methods required a high degree of manual manipulation, and it was not possible to extract ECG waveforms in a fully automatic manner since ECGs generated from different institutions had different printing formats and scanning or quality. Recent advances in object detection and image segmental techniques based on artificial neural networks have made it possible to achieve fully automatic waveform extraction from photos of ECGs taken by a smartphone. To the best of our knowledge, this is the first report of an artificial neural network being used to handle intermediate steps of ECG waveform extraction, including waveform localization, and labelling; gridline and background noise removal; and scale calibration using voltage markers on ECGs. As the overall capabilities of machine learning systems improve, the performance of ECG waveform extraction from photos of ECGs is also expected to improve significantly over time.

Digitheart takes an image as input and outputs a waveform signal in voltage-time format which can then be fed into an ECG diagnostic algorithm. Our study also included a smartphone application as a proof-of-concept device to demonstrate the feasibility of deriving cardiac rhythm diagnosis directly from an ECG photograph.

Our system can act as a platform to facilitate automatic, rapid, cost effective, and large-scale deployment of the state-of-the-art ECG diagnostic algorithms into clinical practice. Smartphone-based ECG diagnostic systems are expected to become more prevalent and accurate in the future if more diagnostic algorithms can be applied.

### Limitations

First, although our system was designed to suit a wide range of ECG formats, the training and validation set was limited to the ECG formats used in Hong Kong public hospitals. With more ECG formats trained by our model, the generalizability of our system can be further enhanced. Second, our AF classifier served only a proof-of-concept purpose to illustrate the possibility of deriving cardiac rhythm from a smartphone-acquired ECG photograph. It is necessary to further develop or incorporate other disease classifiers into the system to realize its full potential.

## CONCLUSION

Using computer vision and machine learning techniques to detect cardiac conditions from photos of ECGs taken with smartphones is feasible. This platform can enable widespread deployment of the latest machine learning-based ECG diagnostic algorithms.

## Abbreviation

AF: Atrial fibrillation
ECG: Electrocardiography
SR: Sinus rhythm

## Author Contributions

Chun-Ka WONG, Yuk-Ming LAU, Hin-Wai LUI and Chung-Wah SIU conceived and designed the study. Chun-Ka WONG, Yuk-Ming LAU, Hin-Wai LUI, Wai-Fung CHAN, Wing-Chun SAN and Chung-Wah SIU designed and created the software and deep learning systems. Wing-Hon LAI, Yee-Man LAU, Duo HUANG, Mi ZHOU and Yangyang CHENG contributed to data acquisition. Chun-Ka WONG and Chung-Wah SIU reviewed and labelled cardiac rhythm of ECG. Chun-Ka WONG, Yuk-Ming LAU, Hin-Wai LUI, Wai-Fung CHAN, Wing-Chun SAN and Chung-Wah SIU performed data analysis and interpretation. Chun-Ka WONG, Yuk-Ming LAU, Hin-Wai LUI, Wai-Fung CHAN, Wing-Chun SAN and Chung-Wah SIU wrote the first draft of the manuscript. Chung-Wah SIU revised the manuscript critically for important intellectual content. All authors have read and approved the final version of the manuscript to be published.

## Acknowledgement

None.

## Data availability statement

Anonymized data will be available from the corresponding author upon reasonable requests up to 3 years after publication of the article. Raw images of the electrocardiograms (ECGs) could not be transferred to third parties owing to patient privacy concerns.

---

**Video 1.** Proof-of-concept smartphone application demonstrating the feasibility of deriving cardiac rhythm directly from photos of electrocardiograms (ECGs).

